# The Association of COVID-19 Incidence with Sport and Face Mask Use in United States High School Athletes

**DOI:** 10.1101/2021.01.19.21250116

**Authors:** Andrew M. Watson, Kristin Haraldsdottir, Kevin Biese, Leslie Goodavish, Bethany Stevens, Timothy McGuine

## Abstract

**Purpose:** To evaluate the influence of sport characteristics and face mask use on COVID-19 incidence among high school athletes.

**Methods:** Surveys were distributed to high school athletic directors throughout the United States regarding sport re-initiation, COVID-19 cases, and risk reduction procedures in fall 2020.

Separate mixed effects Poisson regression models were developed to evaluate the associations between reported COVID-19 incidence and 1) sport characteristics (contact/non-contact, individual/team, indoor/outdoor) and 2) face mask use while playing (yes/no).

**Results:** 991 schools had restarted fall sports, representing 152,484 athletes on 5,854 teams. 2,565 cases of COVID-19 were reported, representing a case rate of 1,682 cases per 100,000 athletes and an incidence rate of 24.6 cases per 100,000 player-days. COVID-19 incidence was lower among outdoor versus indoor sports (incidence rate ratio [IRR]=0.54, 95% CI=0.49-0.60, p<0.001) and non-contact versus contact sports (IRR=0.78 [0.70-0.87], p<0.001), but not team versus individual sports (IRR=0.96 [0.84-1.1], p=0.49). Face mask use was associated with a decreased incidence in girls’ volleyball (IRR=0.53 [0.37-0.73], p<0.001), boys’ basketball (IRR=0.53 [0.33-0.83], p=0.008) and girls’ basketball (IRR=0.36 [0.19-0.63], p<0.001), and approached statistical significance in football (IRR=0.79 [0.59-1.04], p=0.10) and cheer/dance (IRR=0.75 [0.53-1.03], p=0.081).

**Conclusions:** In this nationwide survey of US high school athletic directors representing 152,484 athletes, lower COVID-19 incidence was independently associated with participation in outdoor versus indoor and non-contact versus contact sports, but not team versus individual sports. Face mask use was associated with decreased COVID-19 incidence among indoor sports, and may be protective among outdoor sports with prolonged close contact between participants.

## INTRODUCTION

In an effort to control the spread of the coronavirus disease 2019 (COVID-19) in spring 2020, restrictions were placed on virtually all facets of American society, including the cancellation of school and interscholastic athletics. Early research suggests that school and sport cancellations during the initial months of the COVID-19 pandemic were associated with significant decreases in physical activity and worsening of depressive symptoms in children and athletes.^3,6,8^ It has been projected that prolonged restriction could contribute significantly to long-term increases in obesity and mental health disorders.^2,7,15^ In a nationwide survey of over 13,000 adolescent athletes in May 2020, 37% reported moderate to severe symptoms of anxiety and 40% reported moderate to severe symptoms of depression.^9^ Together these results suggest that isolation and physical inactivity during COVID-19 restrictions may represent a significant threat to physical and mental health in children and adolescents.

Nonetheless, efforts to promote the benefits of youth sports participation are necessarily balanced against the potential risk of COVID-19 transmission. While high school sports have restarted in many areas of the United States, they remain shut down in other areas, with a lack of specific evidence to guide decision-making. A number of organizations have developed risk reduction protocols in an attempt to mitigate the spread of COVID-19 among youth sport participants, but it is widely recognized that there is very little prior research within sport environments to guide these decisions.^1,5,11,12,17^ The available evidence has been limited to case reports in adult recreational athletes, media reports of infections among adults and adolescents associated with interscholastic athletics, and a single evaluation of youth soccer in a small-group, physically-distanced setting.^4,5,13,18^

Organizations have also attempted to classify sports in terms of the risk of COVID-19 transmission during participation. These recommendations are based on a number of characteristics but differ between organizations, as evidence derived from sport contexts is lacking.^1,11,12,17^ In fact, we are aware of no research which has evaluated the relative risks of COVID-19 among athletes between different sports or between sport characteristics such as indoor versus outdoor, individual versus team or contact versus non-contact.

Similarly, the recommendations regarding face mask use during sport participation differ between public health organizations, and the American Academy of Pediatrics (AAP) recently changed its recommendation to encourage their use among youth athletes during most sport contexts.^1,10,12,17^ Although there is general consensus among the scientific community that face mask use in community settings can reduce transmission of COVID-19, there is no evidence specifically within youth sport environments regarding their efficacy, and whether any benefit differs between sports or sport characteristics. Therefore, the purpose of this study was to determine the associations between COVID-19 risk and specific sports, sport characteristics, and face mask use among US high school athletes.

## METHODS

### Study Design

All procedures performed in this study were approved by the Institutional Review Board of the University of Wisconsin-Madison. In collaboration with the National Federation of State High School Associations (NFHS), surveys were distributed to all state high school athletic associations in the United States between November 1, 2020 and November 3, 2020. Among states in which fall high school athletics had restarted, surveys were forwarded on to the athletic directors of high schools within the state. In addition to school name and location, athletic directors were asked whether they had restarted participation in sports since the initial COVID-19 restrictions in the spring of 2020. Those schools who reported reinitiating sports were asked to provide the specific sports and the date of restarting, number of athletes, number of practices and games, number of COVID-19 cases among athletes, and reported sources of infection (if known) within each sport during the months of August, September and October 2020. Schools were asked about their type of instruction during the fall (virtual, in-person) and what restarted sports were using face masks for players while playing. Schools were included if they had any sport that had restarted participation during August, September or October 2020.

### Statistical Analysis

Data were initially evaluated using descriptive statistics, including estimates of central tendency (mean, median) and variability (standard deviation, interquartile range, range) for continuous variables, and counts and percentages for categorical variables. Case rates were expressed as the number of reported cases per 100,000 players (cases / total number of players * 100,000) overall and for each sport. Duration of participation for each sport at each school was determined as the difference in days between the date of restarting and October 31, 2020 and player-days was determined as the product of the number of participating players and duration. Incidence rates were expressed as the number of reported cases per 100,000 player-days (cases / total number of player-days * 100,000) overall and for each sport, with confidence intervals calculated using an exact method.

In addition, the number of cases, total population, case rate and incidence rate during August, September, and October were determined for each state in which a respondent high school was located, from publicly available online information aggregated from the US Centers for Disease Control and local health authorities.^19^ In order to determine whether background state COVID-19 rates were associated with reported COVID-19 rates among high school athletes, the total number of athletes and COVID-19 cases were aggregated by state. For those states with >100 athletes, the relationship between COVID-19 case rates among high school athletes and the general population were evaluated with a linear regression model weighted for the number of players from each state.

For those sports with data from 50 or more schools, the relative risk of each sport was evaluated using a mixed effects Poisson regression model to predict the number of COVID-19 cases for each team with local incidence, instructional delivery type (in-person, virtual), and sport as fixed effects, school as a random effect, and the log of player-days as an offset, yielding an incidence rate ratio (IRR) with Soccer – Boys as the reference (since it represented the median unadjusted incidence rate and is typically considered a moderate risk sport). To evaluate the independent relationships between COVID-19 cases and sport characteristics, a mixed effects Poisson regression model was developed to predict the number of cases with local incidence, instructional delivery type, sport location (indoor, outdoor), sport contact (contact, non-contact), and sport type (individual, team) as fixed effects, school as a random effect and the log of player-days as an offset including data from all sports reported. The specific sports that were reported and their respective characteristics are shown in Supplemental Table 1.

**Table 1.**
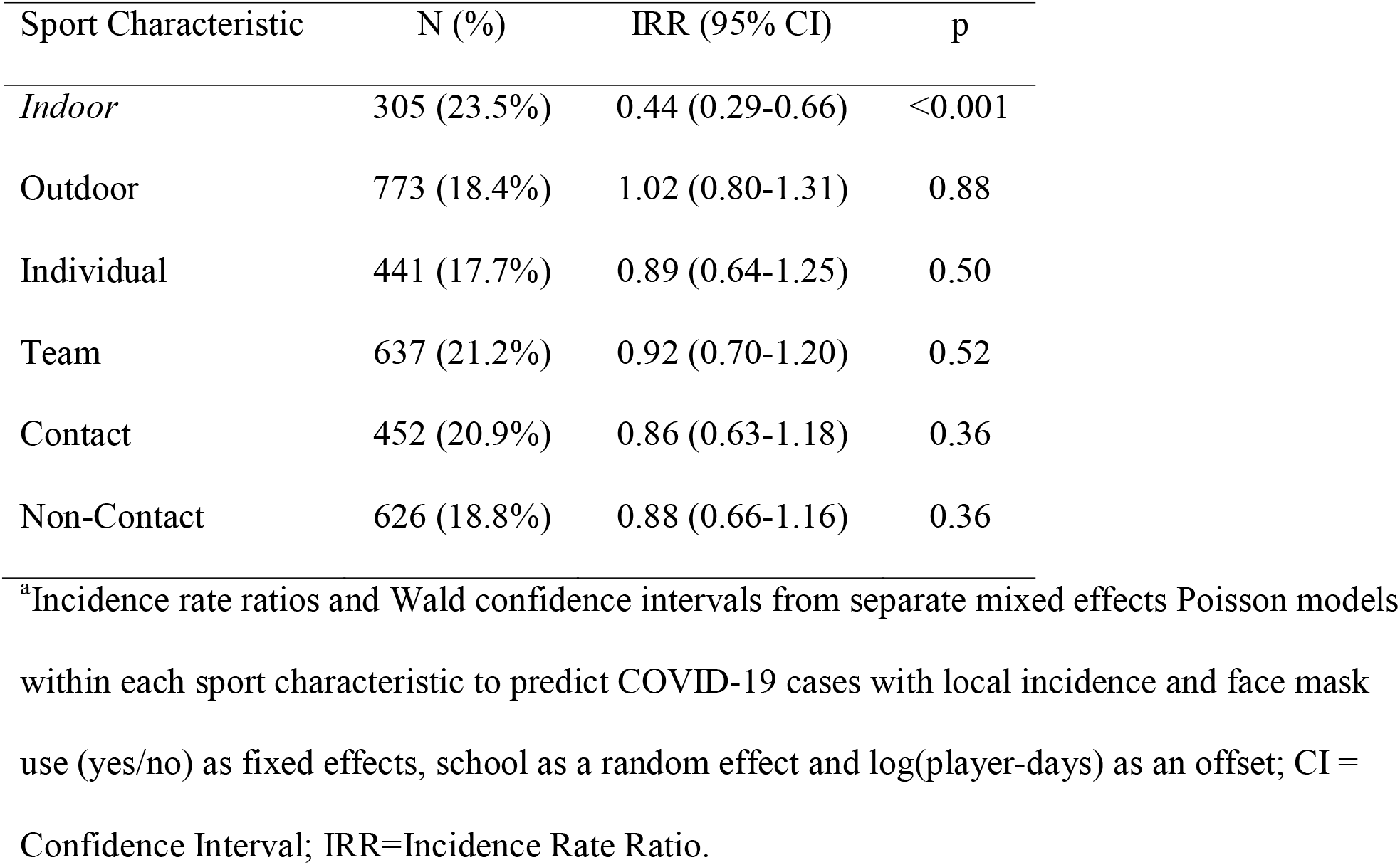
The association of reported face mask use with COVID-19 incidence within each sport characteristic among US high school athletes during fall 2020.^a^

| Sport Characteristic | N (%) | IRR (95% CI) | p |
| --- | --- | --- | --- |
| *Indoor* | 305 (23.5%) | 0.44 (0.29-0.66) | <0.001 |
| Outdoor | 773 (18.4%) | 1.02 (0.80-1.31) | 0.88 |
| Individual | 441 (17.7%) | 0.89 (0.64-1.25) | 0.50 |
| Team | 637 (21.2%) | 0.92 (0.70-1.20) | 0.52 |
| Contact | 452 (20.9%) | 0.86 (0.63-1.18) | 0.36 |
| Non-Contact | 626 (18.8%) | 0.88 (0.66-1.16) | 0.36 |

<sup>a</sup>Incidence rate ratios and Wald confidence intervals from separate mixed effects Poisson models within each sport characteristic to predict COVID-19 cases with local incidence and face mask use (yes/no) as fixed effects, school as a random effect and log(player-days) as an offset; CI = Confidence Interval; IRR=Incidence Rate Ratio.

To evaluate the association between overall COVID-19 incidence and reported face mask use, a mixed effects Poisson regression model was developed to predict the number of cases for each team, with local incidence, instructional delivery type, and face mask use (yes/no) as fixed effects, school as a random effect and the log of player-days as an offset. Similar, separate models were developed for each sport characteristic (indoor, outdoor, contact, non-contact, individual, team) and each specific sport with greater than 40 reported cases. Within each of these sports, incidence rates and 95% confidence intervals were calculated within each sport for those teams reporting face mask use or not and compared descriptively. Swimming was excluded from the face mask analyses. Coefficients from Poisson models were exponentiated to represent IRRs for binary variables and Wald confidence intervals were generated. Significance level was determined *a priori* at the 0.05 level and all tests were 2-tailed. All statistical analyses were performed in R.^14^

## RESULTS

1,508 schools submitted survey responses, of which 991schools had restarted a fall sport. These schools represented 152,484 student-athletes on 5,854 teams that had participated in 159,947 practices and 48,582 games. Eight hundred eighty-nine (89.7%) respondent schools reported utilizing in-person instruction during the fall of 2020. Among the schools that had restarted participation, 2,565 cases of COVID-19 were reported, yielding a case rate of 1,682 cases per 100,000 athletes and an incidence rate of 24.6 (95% CI = 23.7-25.6) cases per 100,000 player-days. Of the cases with a reported, known source, 870 (55%) were attributed to household contact followed by community contact outside sport or school (516, 32%), school contact (115, 7.3%), sport contact (69, 4.3%) and other (24, 1.5%). For those sports with greater than 50 participating schools, the unadjusted COVID-19 incidence rate ranged from 10.4 (Tennis – Boys) to 52.0 cases per 100,000 player-days (Basketball - Girls), as shown in Figure 1 (full data for all sports is available in Supplementary Table 2).

**Table 2.**
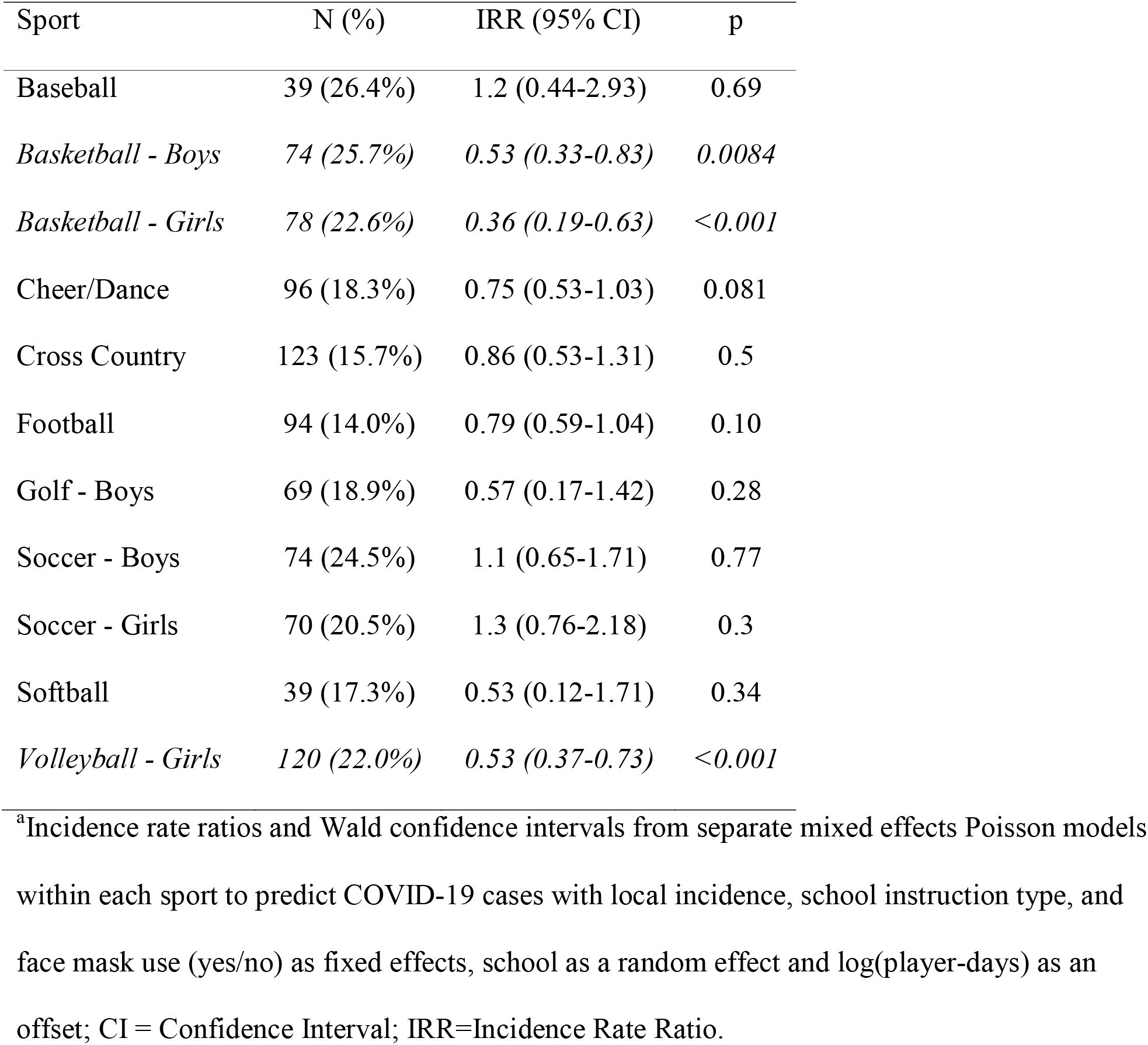
The association of reported face mask use with COVID-19 incidence within each sport among US high school athletes during fall 2020.^a^

| Sport | N (%) | IRR (95% CI) | p |
| --- | --- | --- | --- |
| Baseball | 39 (26.4%) | 1.2 (0.44-2.93) | 0.69 |
| *Basketball - Boys* | *74 (25.7%)* | *0.53 (0.33-0.83)* | *0.0084* |
| *Basketball - Girls* | *78 (22.6%)* | *0.36 (0.19-0.63)* | *<0.001* |
| Cheer/Dance | 96 (18.3%) | 0.75 (0.53-1.03) | 0.081 |
| Cross Country | 123 (15.7%) | 0.86 (0.53-1.31) | 0.5 |
| Football | 94 (14.0%) | 0.79 (0.59-1.04) | 0.10 |
| Golf - Boys | 69 (18.9%) | 0.57 (0.17-1.42) | 0.28 |
| Soccer - Boys | 74 (24.5%) | 1.1 (0.65-1.71) | 0.77 |
| Soccer - Girls | 70 (20.5%) | 1.3 (0.76-2.18) | 0.3 |
| Softball | 39 (17.3%) | 0.53 (0.12-1.71) | 0.34 |
| *Volleyball - Girls* | *120 (22.0%)* | *0.53 (0.37-0.73)* | *<0.001* |

<sup>a</sup>Incidence rate ratios and Wald confidence intervals from separate mixed effects Poisson models within each sport to predict COVID-19 cases with local incidence, school instruction type, and face mask use (yes/no) as fixed effects, school as a random effect and log(player-days) as an offset; CI = Confidence Interval; IRR=Incidence Rate Ratio.

**Figure 1.**
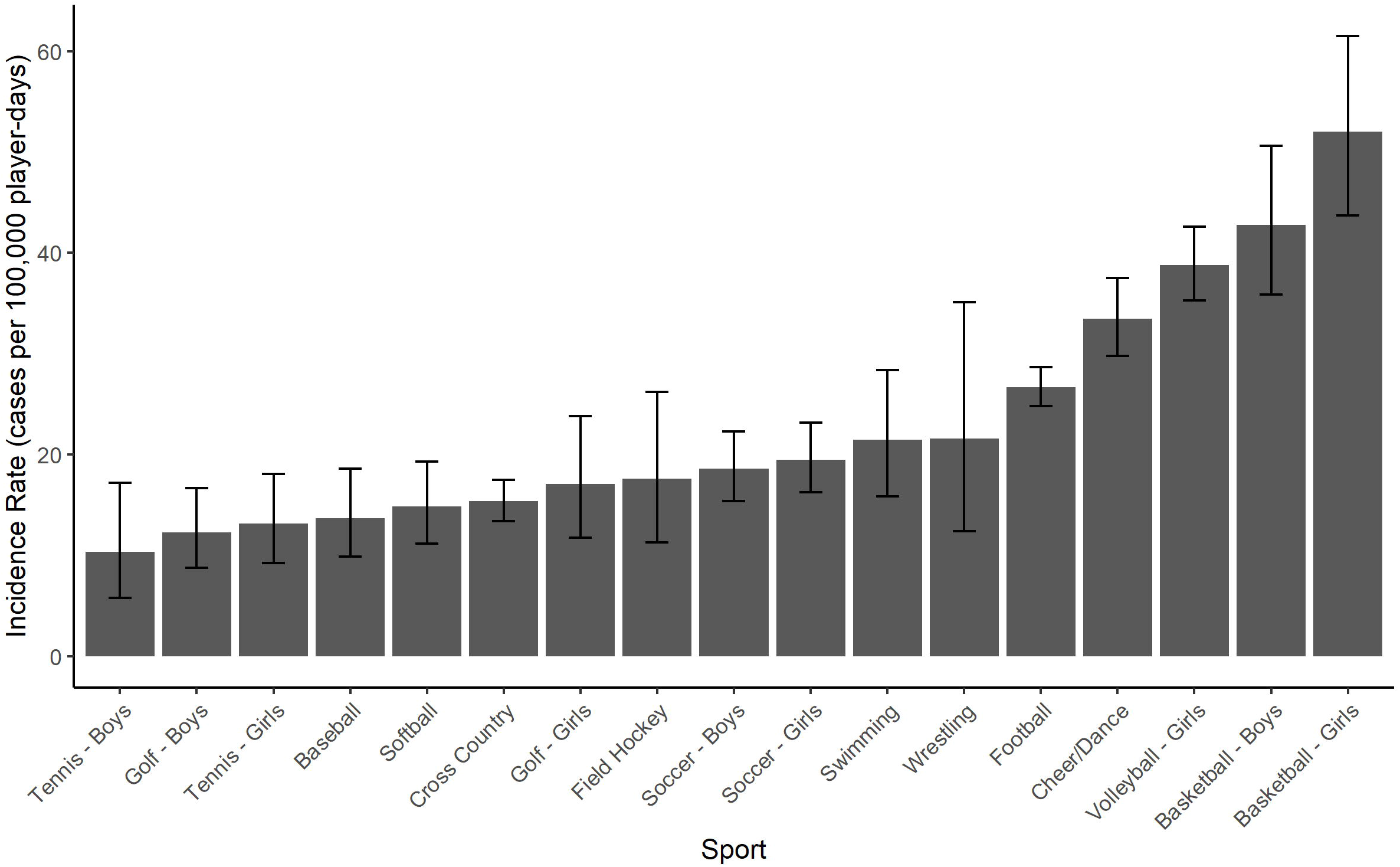
Unadjusted incidence rates of COVID-19 among United States high school sports during fall 2020. Incidence rate is shown as reported cases per 100,000 player-days for those sports with greater than 50 schools reporting re-initiation.

When aggregated by state, the overall COVID-19 case rates for athletes were highly correlated with the case rates for their respective state’s general population (β = 1.09 ± 0.16, r = 0.81, p < 0.001; see Figure 2). The COVID-19 IRRs for specific sports, adjusted for state COVID-19 incidence, instruction delivery type and school repeated measures are shown in Figure 3. After adjusting for state COVID-19 incidence and school instruction type, reported COVID-19 incidence among high school athletes was significantly and independently lower among outdoor versus indoor sports (IRR=0.54 [0.49-0.60], p<0.001) and non-contact versus contact sports (0.78 [0.70-0.87], p<0.001), while no association was identified with respect to team versus individual sports (0.96 [0.84-1.1], p=0.49).

**Figure 2.**
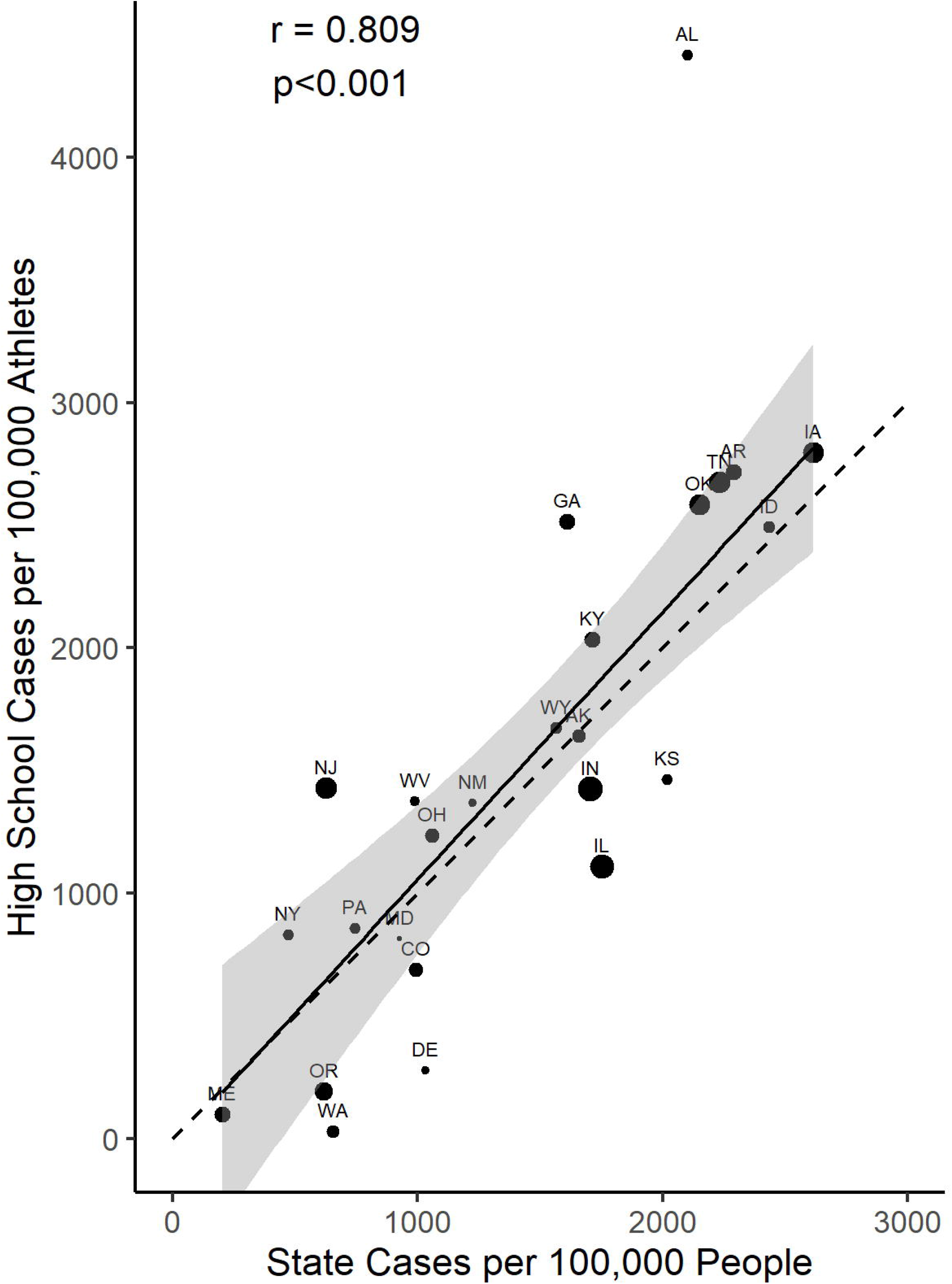
Reported COVID-19 case rates for high school athletes and the general population of their respective states during the fall of 2020. Size of points scaled to number of players from each state and dashed line represents a line of equality. Solid line and shaded area represent regression line and 95% confidence interval from linear model weighted for number of players from each state. r = correlation coefficient.

**Figure 3.**
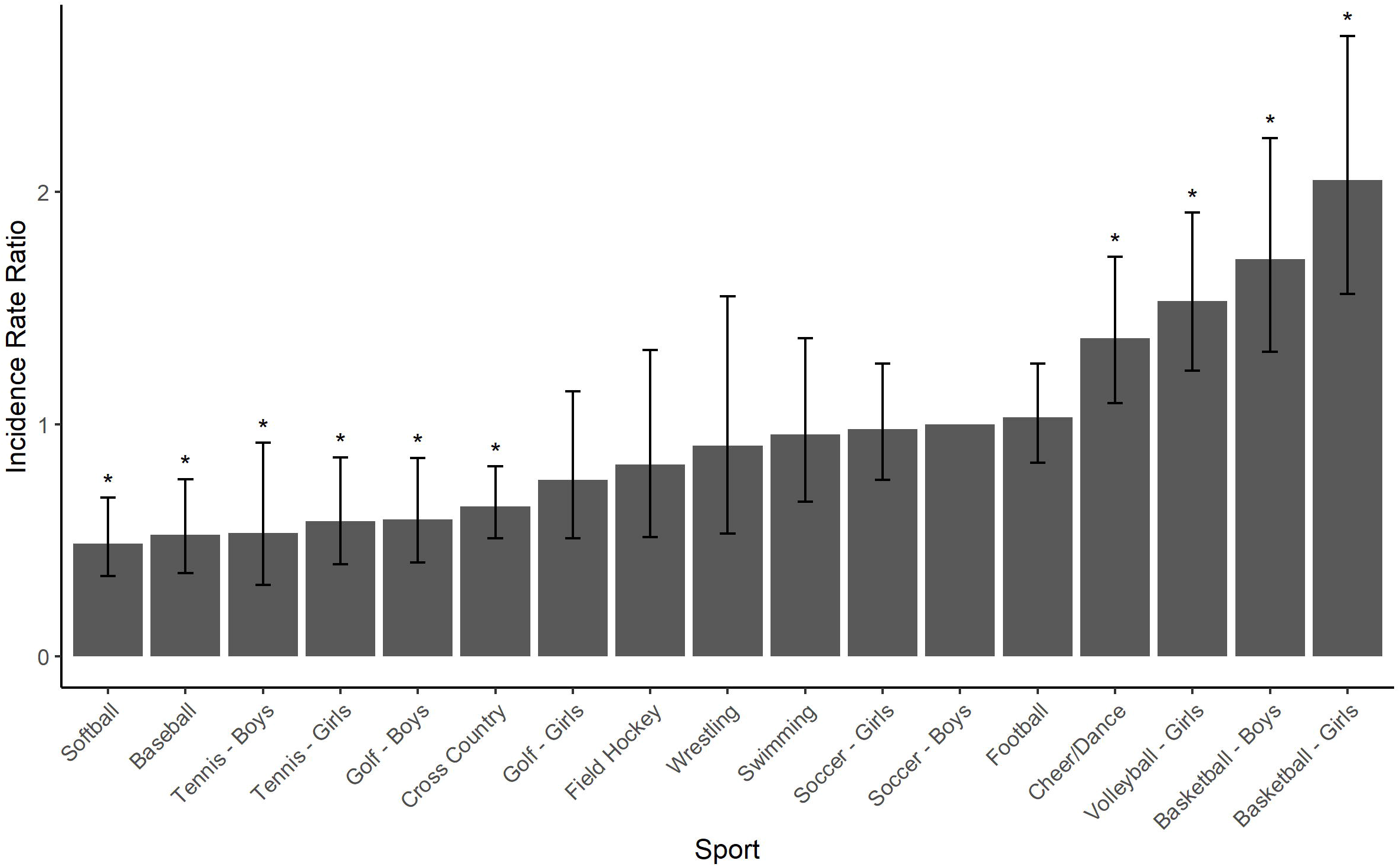
COVID-19 incidence rate ratios during fall 2020 for United States high school sports, adjusted for local (state) COVID-19 incidence, instructional delivery type and repeated measures from the same school. Includes those sports with greater than 50 schools reporting participation, with Soccer – Boys as reference. *p<0.05.

284 schools (28%) reported face mask use by players while playing certain sports, representing 1,677 (28.6%) of all teams participating during the study period. Overall, teams reporting face mask use did not have a lower incidence of COVID-19 among players (IRR = 0.94 [95% CI = 0.75-1.16], p=0.55). However, COVID-19 incidence was lower with face mask use among players participating in indoor sports (Table 1). For those sports with greater than 40 reported cases, differences in COVID-19 incidence between teams with and without face mask use within each sport are shown in Figure 4. Finally, face mask use was associated with a decreased COVID-19 incidence in girls’ volleyball, girls’ basketball, and boys’ basketball, and approached significance in football and cheer/dance, but no association was identified in other sports (Table 2).

**Figure 4.**
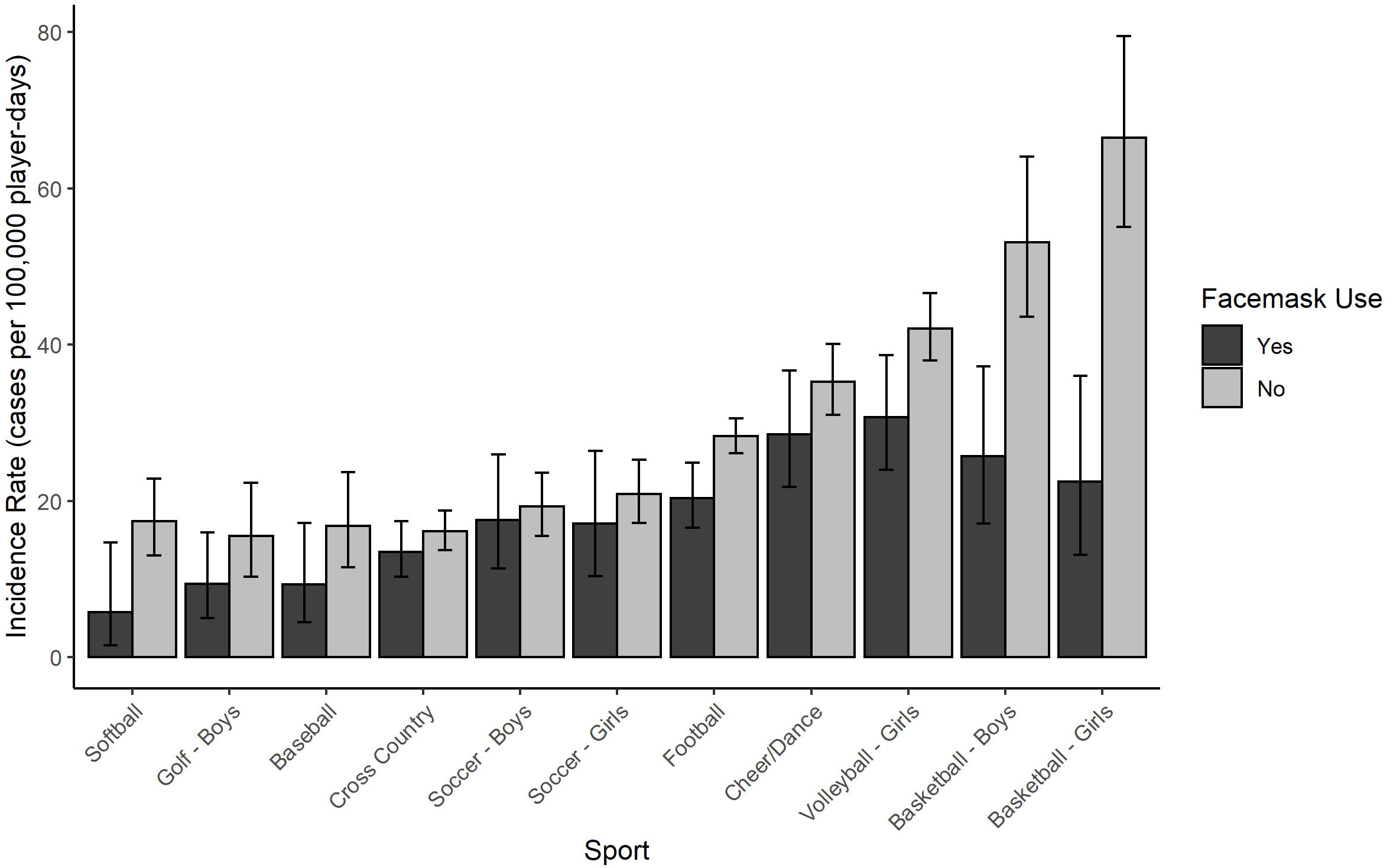
Unadjusted COVID-19 incidence rates reported among US athletes in the fall 2020, comparing teams with or without reported face mask use, within each sport.

## DISCUSSION

These findings suggest that the incidence of COVID-19 among US high school athletes in the fall of 2020 differs between sports and sport characteristics. Although only a small proportion of the cases with a reported source were attributed to sport contact, indoor location and contact were independently associated with an increased incidence rate of COVID-19. This is the first evidence we are aware of that has been derived specifically from a high school sport context, but it is in agreement with prior research and recommendations from various public health organizations that have suggested that COVID-19 is most likely to be transmitted between individuals in close proximity for prolonged periods, and may be more easily transmissible indoors than outdoors.^1,12,17^ Specifically, sport participation indoors versus outdoors appeared to have the strongest influence on COVID-19 incidence within our represented group of athletes, while contact had an independent, yet weaker relationship. We did not find an independent association between COVID-19 incidence and team versus individual sport participation, suggesting that this effect is minimal after accounting for the influences of location and contact.

Public health organizations and sport governing bodies have attempted to classify sports based on expected risk of COVID-19 transmission,^1,11,12,17^ although we are aware of no prior evidence that has evaluated these within sport environments. With respect to the risk categories offered for high school athletics by NFHS,^12^ our findings are in agreement with the suggestion that outdoor, non-contact sports have the lowest COVID-19 incidence. These data also align with the suggestion that sports with close, sustained contact may carry a relatively increased risk, but suggest that indoor location may have the strongest influence on COVID-19 risk. However, it should be noted that wrestling demonstrated an intermediate risk despite being an indoor sport with prolonged, close contact between participants. It is unclear why this would be the case, although the sample size within this sport was relatively small and consequently the confidence intervals were relatively wide, making it difficult to classify the risk associated with this specific sport with confidence.

It should be recognized that not all sports reported participation during the same timeframe and may therefore have had differing background COVID-19 incidence during their respective seasons. Nationwide COVID-19 cases decreased through August and were relatively stable during September, but began increasing in October.^16^ Although we tried to account for differences in local COVID-19 disease burden within our adjusted models by including state COVID-19 rates during the fall months, we cannot exclude the possibility that the higher incidence among traditional winter sports may be partly due to higher local COVID-19 incidence later in the study period when these sports began participation.

We found that face mask use was associated with a decreased incidence of COVID-19 among specific sports. In general, those sports with the highest incidence of COVID-19 were also found to have the greatest benefit from reported face mask use. Specifically, COVID-19 incidence was lower among indoor sports in which face masks were reportedly used when evaluated collectively, but this was also true within volleyball, girls’ basketball, and boys’ basketball when evaluated individually. Importantly, reported COVID-19 incidence among indoor team sports (volleyball, basketball) when using face masks appeared comparable to the incidence among outdoor team sports, suggesting that the increased risk associated with being indoors may be reduced considerably by face mask use. Face mask use also appeared to be associated with a decreased COVID-19 incidence in football and cheer/dance, although this did not reach statistical significance. This may be attributable to the relatively small proportion of the teams in these sports that reported face mask use, and a larger sample of teams using face masks in these sports may have revealed a significant association.

While face mask use was not found to be associated with COVID-19 incidence among other outdoor contact sports, “contact” as a sport characteristic surely exist along a continuum with respect to the time spent in close proximity to other players. The risk of COVID-19 transmission likely varies between sports with brief contact and relatively little time spent within close proximity to others, to sports with prolonged periods of close contact that constitute an increased likelihood of sufficient exposure for COVID-19 transmission between participants. This supports the suggestion that although face mask use did not appear to have a large effect within sports like soccer, it may still be protective in an outdoor sport with sustained close contact.^1^

Face mask use in the community has been widely recommended by public health agencies but remains a contentious issue within the public at large. Within sports, recommendations differ between organizations, and recently the AAP revised their recommendations regarding face mask use in youth sports, suggesting that they be used in most sports contexts when it is safe to do so.^1^ These differences likely represent the fact that there has been no primary evidence regarding the utility of face masks to reduce COVID-19 transmission during sport participation. While we cannot directly evaluate the true transmission rate within the data available, our findings nonetheless suggest that face masks may have a meaningful influence on COVID-19 risk among indoor sports and outdoor sports with sustained close contact, but less influence among outdoor sports with less time spent in close proximity to other players.

Although we were unable to identify publicly available, state-specific, adolescent case rates during the fall months for many of the represented states within our sample, we nonetheless identified a strong relationship between reported COVID-19 case rates in our high school athletes and the COVID-19 case rates among the general population in their respective states. In addition, the majority of cases with a reported source were attributed to household and community contact with a much smaller proportion attributed to school or sport contact. This may suggest that COVID-19 incidence among adolescent athletes is largely reflective of background COVID-19 rates within their community. The overwhelming majority of schools reported in-person instruction, making it difficult to fully evaluate the role of in-person school instruction in COVID-19 incidence among high school athletes. Nonetheless, we included school instruction type within our adjusted models in order to account for the potential confounding role this could play in comparing different groups. Importantly, it should be recognized that this study cannot comment on the incidence or transmission risk of COVID-19 among attendees at high school sporting events such as fans, coaches, staff, and spectators. While this risk remains undefined, it nonetheless represents a potential contribution to community COVID-19 spread and risk mitigation procedures should continue to be prioritized to not only protect athletes but also to help reduce the risk of infection among attendees.

This study has several limitations. The information is self-reported by the athletic directors of each school and cannot be directly verified through medical records or another independent source. Local, state-level daily COVID-19 case data was often not available for adolescents or children, so our adjusted models could only account for the population-level background incidence from each state. Nonetheless, we found that reported case rates from our sample and the case rates from the state general populations were highly related. As mentioned above, the incidence of COVID-19 was likely not stable throughout the fall in many areas, and those sports that initiated play during periods of increased local incidence (winter sports in October, for example) may be biased toward a higher incidence that is not directly attributable to the sport itself. Reported sources of infection were provided by the schools themselves and not through formal contact tracing by local health authorities. We cannot directly account for the possibility of transmission between players that went unidentified. Finally, while this data represents information regarding a large number of male and female high school athletes from a nationwide sample, it may not be generalizable to other populations.

In conclusion, this study suggests that certain high school sports and sport characteristics may have a greater relative risk of COVID-19 and that face mask use may help reduce the risk of COVID-19 among adolescent athletes in sports with higher risk. Specifically, indoor sports appear to have a greater risk of COVID-19 infection among participants, while outdoor, non-contact sports have the lowest risk. However, face mask utilization was associated with a significantly decreased incidence of COVID-19 in indoor sports, and this appeared to mitigate a large portion of the increased risk. Given the general lack of information regarding COVID-19 risk among youth sport participants, this information may help guide decision-making to reduce the risk of COVID-19 transmission, while facilitating the wide-ranging physical and mental health benefits of sport participation.

## Data Availability

Data is available upon reasonable request.

