## Supplemental Table 1 for "The Association of COVID-19 Incidence with Sport and Face Mask Use in United States High School Athletes"

Supplemental Table 1. Characteristics of the participating sports from US High Schools in fall 2020.

| Sport | Indoor/Outdoor | Contact/Non-Contact | Individual/Team |
| --- | --- | --- | --- |
| Baseball | Outdoor | Non-Contact | Team |
| Basketball - Boys | Indoor | Contact | Team |
| Basketball - Girls | Indoor | Contact | Team |
| Cheer/Dance | Outdoor | Non-Contact | Individual |
| Cross Country | Outdoor | Non-Contact | Individual |
| Field Hockey | Outdoor | Contact | Team |
| Football | Outdoor | Contact | Team |
| Golf - Boys | Outdoor | Non-Contact | Individual |
| Golf - Girls | Outdoor | Non-Contact | Individual |
| Gymnastics | Indoor | Non-Contact | Individual |
| Hockey - Boys | Indoor | Contact | Team |
| Hockey - Girls | Indoor | Contact | Team |
| Lacrosse - Boys | Outdoor | Contact | Team |
| Lacrosse - Girls | Outdoor | Contact | Team |
| Rugby | Outdoor | Contact | Team |
| Soccer - Boys | Outdoor | Contact | Team |
| Soccer - Girls | Outdoor | Contact | Team |
| Softball | Outdoor | Non-Contact | Team |
| Swimming | Indoor | Non-Contact | Individual |
| Tennis - Boys | Outdoor | Non-Contact | Individual |
| Tennis - Girls | Outdoor | Non-Contact | Individual |
| Track and Field | Outdoor | Non-Contact | Individual |
| Volleyball - Boys | Indoor | Non-Contact | Team |
| Volleyball - Girls | Indoor | Non-Contact | Team |
| Wrestling | Indoor | Contact | Individual |
