## Supplemental Table 2 for "The Association of COVID-19 Incidence with Sport and Face Mask Use in United States High School Athletes"

Supplementary Table 2. The number of participating schools, players, reported cases, player-days, COVID-19 case and incidence rates by sport among United States high schools in the fall of 2020.

|  | Schools | Players | Cases | Player-Days | Case Rate^a^ | Incidence Rate^b^ |
| --- | --- | --- | --- | --- | --- | --- |
| Baseball | 154 | 4111 | 42 | 305933 | 1022 | 13.7 (9.9-18.6) |
| Basketball - Boys | 295 | 7251 | 137 | 319997 | 1889 | 42.8 (35.9-50.6) |
| Basketball - Girls | 353 | 6717 | 137 | 263451 | 2040 | 52 (43.7-61.5) |
| Cheer/Dance | 537 | 12457 | 301 | 898521 | 2416 | 33.5 (29.8-37.5) |
| Cross Country | 800 | 19846 | 226 | 1470503 | 1139 | 15.4 (13.4-17.5) |
| Field Hockey | 90 | 2718 | 24 | 136396 | 883 | 17.6 (11.3-26.2) |
| Football | 687 | 37184 | 742 | 2776575 | 1995 | 26.7 (24.8-28.7) |
| Golf - Boys | 382 | 4342 | 41 | 333568 | 944 | 12.3 (8.8-16.7) |
| Golf - Girls | 379 | 2449 | 34 | 199370 | 1388 | 17.1 (11.8-23.8) |
| Gymnastics | 8 | 92 | 0 | 6531 | 0 | 0 (0-56.5) |
| Hockey - Boys | 9 | 200 | 5 | 10487 | 2500 | 47.7 (15.5-111) |
| Hockey - Girls | 2 | 25 | 0 | 1680 | 0 | 0 (0-220) |
| Lacrosse - Boys | 17 | 595 | 2 | 27283 | 336 | 7.33 (0.89-26.5) |
| Lacrosse - Girls | 14 | 463 | 0 | 23493 | 0 | 0 (0-15.7) |
| Rugby | 1 | 40 | 2 | 2440 | 5000 | 82 (9.9-296) |
| Soccer - Boys | 311 | 10439 | 119 | 639473 | 1140 | 18.6 (15.4-22.3) |
| Soccer - Girls | 354 | 10127 | 129 | 660217 | 1274 | 19.5 (16.3-23.2) |
| Softball | 234 | 4383 | 55 | 370140 | 1255 | 14.9 (11.2-19.3) |
| Swimming | 173 | 4259 | 49 | 227946 | 1151 | 21.5 (15.9-28.4) |
| Tennis - Boys | 134 | 1861 | 15 | 143635 | 806 | 10.4 (5.8-17.2) |
| Tennis - Girls | 203 | 4148 | 37 | 281075 | 892 | 13.2 (9.3-18.1) |
| Track and Field | 48 | 1544 | 0 | 66856 | 0 | 0 (0-5.52) |
| Volleyball - Boys | 11 | 169 | 0 | 9938 | 0 | 0 (0-37.1) |
| Volleyball - Girls | 566 | 15075 | 452 | 1164004 | 2998 | 38.8 (35.3-42.6) |
| Wrestling | 92 | 1989 | 16 | 73956 | 804 | 21.6 (12.4-35.1) |
| Total | 5854 | 152484 | 2565 | 10413468 | 1680 | 24.6 (23.7-25.6) |

^a^Case rates shown as cases per 100,000 players. ^b^Incidence rates shown as cases per 100,000 player-days with exact 95% confidence intervals.
